# Estimating Blood Pressure from the Electrocardiogram: Findings of a Large-Scale Negative Results Study

**DOI:** 10.1101/2025.07.25.25332198

**Authors:** Seyedeh Somayyeh Mousavi, Sajjad Karimi, Mohammad Sina Hassannia, Zuzana Koscova, Ali Bahrami Rad, David Albert, Gari D. Clifford, Reza Sameni

## Abstract

**Objective:** Electrocardiography and blood pressure (BP) measurement are two widely used tools for diagnosis and monitoring cardiovascular diseases. While the electrocardiogram (ECG) and BP have been considered complementary modalities, there are also systematic relationships between them. Therefore, advancements in portable and wearable ECG devices, along with promising results in cuff-less BP measurement using a combination of ECG and other bio-signals have led researchers to hypothesize the possibility of estimating BP using only ECG. However, the literature is divided on this topic: some studies support this hypothesis, while others reject it.

**Approach:** In this study, machine-learning (ML) models were developed to explore this hypothesis by estimating BP from 30-second ECGs using an extensive dataset from AliveCor Inc., which includes 124,427 records from 7,412 subjects. The ECG and BP recordings were asynchronous with variable counts and time lags. Therefore, a 3.5-minute time window before and after each ECG recording was used to calculate the mean BP measurement.

**Main Results:** Sex-aware ML models were trained using a comprehensive feature vector comprising 280 features: 128 explainable ECG features developed by the research team and 150 ECG features extracted by the Black Swan team, one of the top-performing teams in the PhysioNet Challenge 2017. Additionally, the average time gap between each ECG and the corresponding BP measurement, along with the subject’s age, were included as two supplementary features.

**Significance:** Our best ML models achieved a mean absolute error (MAE) of 12.59 mmHg for systolic blood pressure (SBP) and 7.43 mmHg for diastolic blood pressure (DBP), with correlation coefficients of 0.35 and 0.38 between the predicted and actual values, respectively. Therefore, the results indicate no significant relationship between BP and ECG. In conclusion, ML models cannot correctly estimate BP values from ECGs, rejecting the proposed hypothesis.

## 1. Introduction

Blood pressure (BP) measurement and electrocardiography are two complementary methods widely used for cardiovascular monitoring and diagnosis. BP is influenced by cardiac mechanical function and systemic vascular resistance [1]. When the heart contracts, it creates a pulsatile pressure wave in the arterial system [2]. The systolic blood pressure (SBP) represents the maximum and the diastolic blood pressure (DBP) reflects the minimum of the pressure wave in each cardiac cycle, both of which are time-varying due to natural fluctuations and measurement errors and biases, such as those caused by respiration [1, 3, 4]. Both tonic and cyclic fluctuations in the blood pressure wave provide critical insights into cardiovascular health, making BP monitoring a standard in patient care and an effective tool for cardiovascular diseases (CVDs) diagnosis and management [1, 5]. The guidelines for hypertension recommend that symptomatic individuals regularly monitor their BP [6].

On the other hand, the Electrocardiogram (ECG) measures the electrical function of the heart and captures the electro-physiological patterns of depolarization and re-polarization during each cardiac cycle [7, 8]. ECG recording is cost-effective, accurate and commonly available in most health centers and outside clinical settings using portable and wearable ECG monitors, making it suitable for long-term cardiac monitoring and CVD detection [9–11].

Previous studies have attempted to estimate BP using machine-learning (ML) or deep learning (DL) methods from only photoplethysmography (PPG) signals [12, 13], or from a combination of PPG with other biosignals such as the output signal of a Hall sensor [14, 15], the modulated magnetic signature of the blood [16], ballistocardiography [17, 18], and impedance plethysmography [19, 20]. Researchers have further hypothesized the feasibility of estimating BP using only ECGs and electro-physiological features [21–24]. Rapid advancements in home care devices, such as portable ECG devices, smartwatches, and smartphones, have further motivated efforts to integrate BP measurement functionality into these technologies [9]. However, the literature is divided on whether BP can be accurately estimated from ECGs: those whose results support this hypothesis and those that do not.

In 2008, Ali Hassan et al. [25] developed a linear regression model to estimate SBP from heart rate (HR) extracted from 30-second ECG recordings of 10 normal-ECG subjects. BP values were also measured manually. For each individual, 20 records were used to develop the regression model and the 10 remaining ones were used for testing. To generalize SBP estimation for new subjects, the final model slope was obtained by averaging the individual regression slopes across all participants.

In 2018, Simjanoska et al. [26] developed an ML model for estimating BP using ECGs. The study analyzed 3,129 ECGs with a length of 30 seconds, from 51 subjects, including both healthy and unhealthy individuals. The feature vector consisted of seven components: signal mobility, signal complexity, fractal dimension, entropy, autocorrelation, age, and hypertension classification. Four models were developed: one classification model to predict the hypertension group, and three regression models to estimate SBP, DBP, and Mean Arterial Pressure (MAP). The mean absolute error (MAE) and standard deviation (SD) of the regression model were 7.72*±*10.22 mmHg for SBP and 9.45*±*10.03 mmHg for DBP. The group further extended their study by employing a different pre-processing approach, adjusting the cutoff frequency of filters, and utilizing ECGs with different lengths of 10, 20, and 30 seconds [27]. The results showed an MAE and SD of 16.60*±*11.05 and 9.24*±*7.85 mmHg for SBP and DBP, respectively.

In 2020, Miao et al. [28] developed a DL model for estimating BP from ECG, utilizing a residual network (ResNet) and long short-term memory (LSTM), to capture both time and spatial information from ECGs. The model was trained and tested on the public Multiparameter Intelligent Monitoring in Intensive Care (MIMIC-III) database, which includes ECG and invasive BP information from individuals in critical care units (CCUs) [29]. The pre-processed dataset consisted of 1,711 subjects and 897,743 records, each with a length of 2.5 seconds. The developed approach achieved a ME with a SD of *−*0.22*±*5.82 mmHg for SBP and *−*0.75*±*5.62 mmHg for DBP. The correlation coefficients between the estimated and actual values were 0.88 and 0.71 for SBP and DBP, respectively. Table 1 presents the results of studies based on ML and DL approaches for estimating BP using ECGs.

**Table 1.**
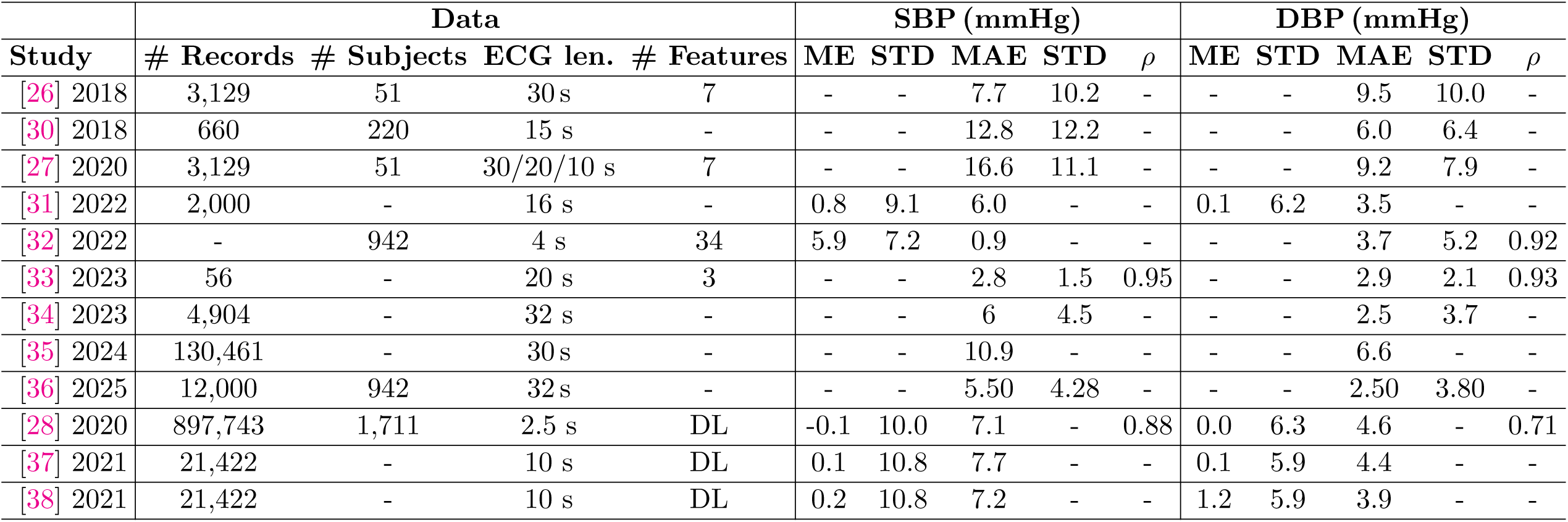
Comparison of studies using machine learning and deep learning (DL) models for blood pressure estimation based solely on ECG data.

At the same time, some research has questioned the feasibility of ECG-based BP estimation. Sato et al. [23] and Mukkamala et al. [24] are two studies in this category, which based on the electrophysiology of BP and ECG and the shortcomings in the reported results in the literature, have debated that accurate ECG-based BP estimation is unfeasible. They have not conducted any independent experiments to support this claim.

This study aims to explore the feasibility of estimating BP using only ECGs using ML models trained on a large ambulatory dataset, while addressing shortcomings in former methodologies. A comprehensive set of 278 engineered features, derived from the time, frequency, and time-frequency domains of the ECGs, and used as inputs to regression models for BP estimation. The models are designed to be demographic-aware by incorporating the sex and age of subjects, which are known to significantly influence BP values [1, 39]. All ECGs are standardized to a fixed length of 30 seconds to ensure consistency across records. Detailed data cleaning, sub-sampling, and standard cross-validation techniques are used to ensure that the results are not biased. *Our findings most strongly support studies that have concluded accurate ECG-based blood pressure estimation is unfeasible*.

## 2. Method

### 2.1. Database

The data used in this study consists of ECG and BP measurements from two databases collected over two years from August 2019 to March 2021 by AliveCor (Mountain View, CA, USA), using the following devices:

i. OMRON Complete (Omron Healthcare, Kyoto, Japan), which is an integrated BP monitor and single-lead ECG;
ii. KardiaMobile (AliveCor, Mountain View, CA, USA) for collecting single-lead ECGs and independent BP readings from portable BP devices (Omron Healthcare, Kyoto, Japan).

To note, the ECG and BP were self-recorded asynchronously in non-clinical settings, with variable numbers of BP and ECG per subject and varying time gaps between the two modalities (varying between seconds and hours). The ECG dataset comprises 180,790 records from 10,624 subjects, with a minimum time gap of 30 seconds between two consecutive ECG recordings for each unique subject. ECGs were recorded at a sampling frequency of 300 Hz. The BP dataset consists of 21,227,729 measurements, corresponding to 297,965 subjects. A total of 10,346 subjects, which were common between the ECG and BP datasets, were shortlisted for this study.

### 2.2. Data cleaning

The data cleaning process is summarized in Fig. 1. Accordingly, records were selected from the matched dataset based on the following criteria:

i. The analysis was limited to adult male and female subjects aged between 18 and 90 years at the time of ECG recording. Subjects with unknown sex or with age outside [18, 90] were excluded from the analysis.
ii. Records with misreported DBP values higher than SBP were removed. Then, thresholds were applied to define valid BP ranges. Valid blood pressure ranges were set to DBP between 20–200 mmHg and SBP between 30–300 mmHg. These thresholds are consistent with the pre-processing approach used in our previous study, which analyzed approximately 75 million BP values from the general population [39].
iii. The dataset included ECG classification labels generated by AliveCor’s proprietary ECG analysis software, which labels signals as “sinus rhythm”, “atrial fibrillation”, “bradycardia”, “tachycardia”, “unclassified”, “too short”, “unreadable” and missing values. Records labeled as “unreadable” or with missing labels were removed.
iv. For consistency, ECG record lengths were fixed to 30 s, and the records shorter than 30 s were excluded from the analysis. Previous studies indicate that this duration is sufficient for capturing essential ECG features, especially for rhythm analysis and heart rate variability [40]. 98% of the ECG database complied with this requirement. For consistency, the ECGs longer than 30 s were truncated to the first 30 s.
v. The ECG and BP data were collected asynchronously, resulting in varying time gaps between the ECG and BP measurements of the same subject. Given that both signals naturally fluctuate over time, we defined a maximum allowable time gap such that BP variability within this window would be minimal—ensuring that estimating BP from ECG remained both meaningful and clinically relevant. To determine this threshold objectively, we referred to acceptable BP error margins from BP device standards and reported rates of short-term BP variability in the literature. Presumably, as long as the time-gap between ECG and BP collection is within these thresholds, any BP change during that interval would fall within an acceptable error margin—making the ECG-BP pairing valid for estimation purposes. According to the Association for the Advancement of Medical Instrumentation (AAMI) standard, the mean BP error in BP measurement devices should be less than 5 mmHg [41]. To identify the time window during which a 5 mmHg change in BP might occur, relevant literature was reviewed. Most studies reported mean BP differences over 30-minute or longer intervals [42–48]. From these studies, reported mean BP differences and their corresponding time windows were extracted to estimate the “rate of BP variation” over time. Using these rates, the time intervals corresponding to the negligible 5 mmHg change were calculated by dividing 5 mmHg by the rate of change. The resulting estimates ranged from 3.5 to 38 minutes. The minimum value (3.5 minutes), was considered as the acceptable short-term window, which we considered as the maximum allowable time gap between ECG and BP recordings.
vi. Many subjects had multiple BP measurements within the acceptable BP-ECG time interval window. For each subject and ECG, all BP measurements within the acceptable time window of (3.5 minutes) were averaged. Averaging BP measurements within short time windows is a standard procedure in clinical practice, which results in more accurate BP measurements [1], and reduction of measurement biases.

**Figure 1.**
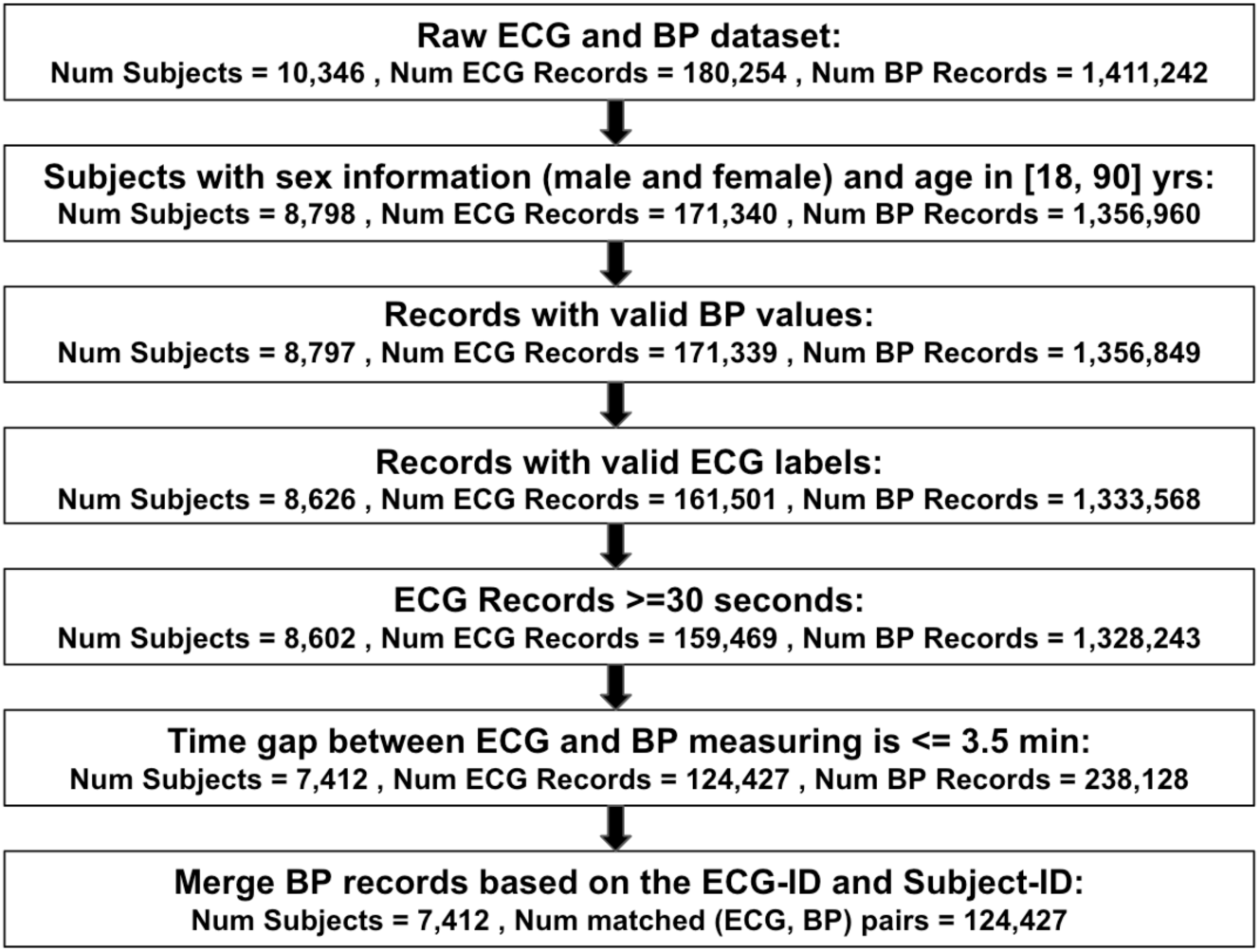
Data cleaning and workflow process for developing machine learning models to estimate blood pressure using only ECGs. Abbreviations: valid label: “sinus rhythm”, “atrial fibrillation”, “bradycardia”, “tachycardia”, “too short”, “unclassified”, invalid label: “unreadable” and missing values. Additionally, in the final stage up to 6 records per subject were selected, based on the median number of records per subject after excluding an outlier with 3500 records and subjects with only one record.

The final filtered dataset included 124,427 pairs of BP and ECG records from 7,412 subjects. Table 2 summarizes the distribution of the final dataset by sex and ECG labels, where “Normal” represents “sinus rhythm” and all other labels are considered “Abnormal”. Also, Table 3 presents the statistical distribution of BP datasets by sex and ECG labels, and Fig. 2 illustrates the 95% percentile range contours for males and females BP distribution. The mean SBP and DBP for each group are marked with dots.

**Table 2.** Distribution of the final processed ECG records based on sex and label.

| Sex | # Total | # Abnormal | # Normal |
| --- | --- | --- | --- |
| Female | 23,597 | 2,846 | 20,751 |
| Male | 100,830 | 14,462 | 86,368 |

**Table 3.** Minimum, maximum, mean, and standard deviation of systolic and diastolic blood pressure (SBP and DBP, in mmHg) of the filtered dataset, categorized by sex and ECG label, for all records and those with sinus rhythm (Normal)

| Sex | BP type | ECG label | Min | Max | Mean | STD |
| --- | --- | --- | --- | --- | --- | --- |
| Male | SBP | All | 60 | 227 | 127.1 | 16.0 |
| Male | SBP | Normal | 65 | 227 | 127.3 | 15.8 |
| Male | DBP | All | 40 | 165 | 80.2 | 10.5 |
| Male | DBP | Normal | 40 | 165 | 80.1 | 10.2 |
| Female | SBP | All | 61 | 215 | 123.8 | 17.0 |
| Female | SBP | Normal | 62 | 215 | 124.0 | 16.7 |
| Female | DBP | All | 40 | 147 | 78.9 | 11.0 |
| Female | DBP | Normal | 40 | 147 | 78.8 | 10.8 |

**Figure 2.**
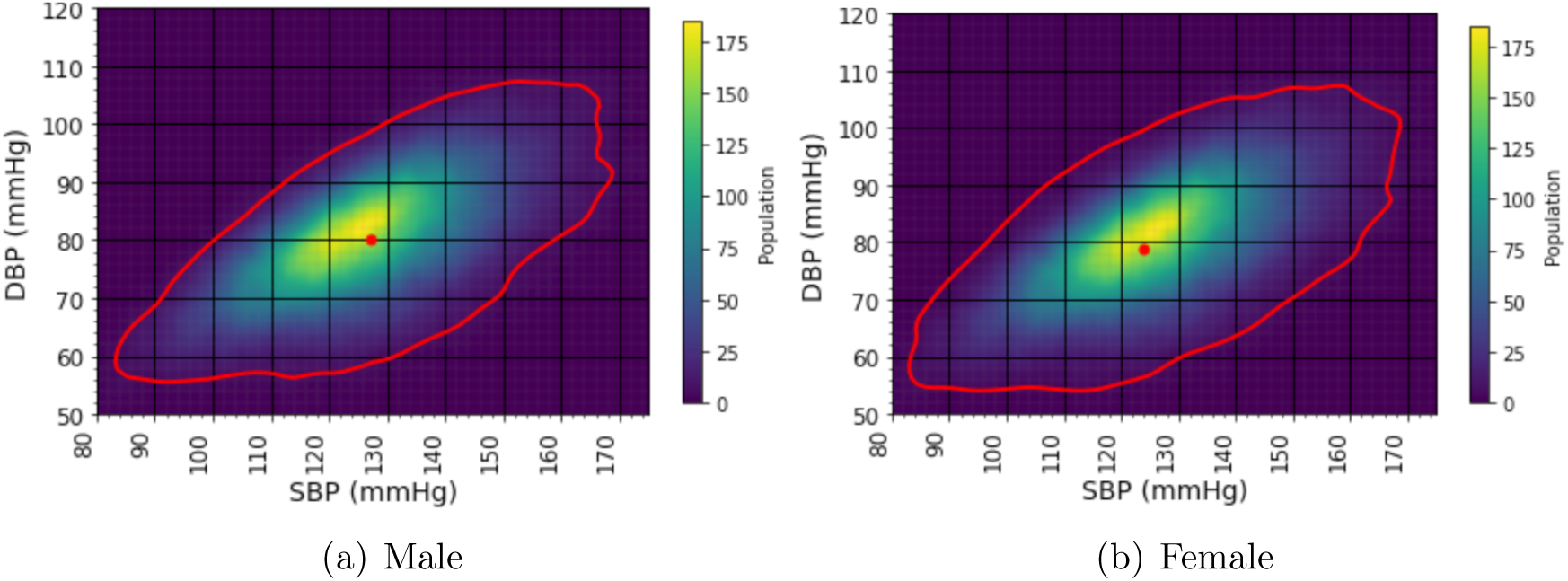
Comparisons of blood pressure distributions between sexes in the pre-processed data presented through heatmaps and contour plots, representing the 95% percentiles of the BP values within the contours. Dots indicate the mean SBP and DBP values. Mean SBP values are 127.1 and 123.8 mmHg, and mean DBP values are 80.2 and 78.9 mmHg for male and female subjects, respectively.

### 2.3. ECG pre-processing

The ECG records were band-pass filtered with a band-pass frequency between 0.1 Hz and 100 Hz, and a notch filter at 50 or 60 Hz, depending on the local power line frequency.

The notch filter was designed using a second-order infinite impulse response (IIR) filter (iirnotch in MATLAB) with a quality factor (Q) of 40 and was applied.

### 2.4. Feature extraction

A total of 280 features were extracted from each ECG record, including: 128 interpretable features developed by our team; 150 features extracted by the Black Swan team [49]; the time gap between the ECG and the average time of the corresponding BP measurements (within the 3.5-minute time windows); and the subject age. To enable the replication of the implemented process, the complete feature set is described below.

i. *Beat signal-to-noise ratio*: To quantify beat-to-beat morphological consistency in the ECG over the 30 s segment, a signal-to-noise ratio (SNR) index was computed and assigned to each beat. R-peaks were first detected using the OSET robust R-peak detector function peak det likelihood [50], and individual beats were segmented using a window of *W* samples centered around each R-peak. Robust weighted average (RWA) and robust beat median (RBM) beats were then calculated, following the method in [51]. For each beat, the residual was computed as the difference from the RWA or RBM beat, and the beat SNR was defined as the power ratio between the original beat and the mean/median-based residuals. These SNRs capture both morphological deviations and measurement noise.
ii. *Heart Rate Variability and Heart Rate Metrics:* After ECG R-peak detection, R-R intervals were computed and converted to instantaneous heart rate (HR) values in beats per minute (bpm). The HR sequence was next summarized using the mean, median, 5th percentile, and 95th percentile. Heart rate variability (HRV) was assessed using the standard deviation of R-R intervals (SDNN) and the root mean square of successive differences (RMSSD) [52].
iii. *Time Interval Measurements*: Fiducial points for each beat were extracted using the fiducial det lsim function from OSET [50]. Using these points, key ECG time intervals were calculated, including the QRS complex duration, QT interval, PR interval, ST interval, PR segment level, and ST segment level. Additional intervals were computed between specific peak pairs: P-R, Q-R, S-R, and T–R, to capture more detailed temporal relationships between waveform components. Corrected QT intervals (QTc) were also derived using the Bazett (QTc-B) and Fridericia (QTc-F) corrections [53].
iv. *Amplitude and Morphological Area Metrics*: Amplitude and area-based features were computed using fiducial points marking the onset, peak, and offset of each ECG waveform component. For each component, the amplitude and the area under the curve (sum of ECG values from onset to offset) were calculated. In addition, we computed the amplitude ratio of the R peak to other major peaks (P, Q, S, and T), and the amplitude difference between the S and T peaks across the ST segment.
v. *Amplitude-to-Timing Ratios*: For each beat, the difference between the R-peak amplitude and the amplitude of other peaks was divided by the time interval between the R peak and the corresponding peak, providing a measure of waveform shape (slope).
vi. *Signal Mobility and Complexity* : *Mobility* was computed as the ratio of the variance of the first derivative of the ECG to the variance of the ECG [26, 27, 54]. *Complexity* was calculated as the ratio of the variance of the second derivative to the variance of the first derivative, divided by the mobility value [26, 27, 54].
vii. *Singular Value Decomposition Metrics*: Singular value decomposition (SVD) has been shown to encode ECG beat variability [55]. ECG beats were segmented around each R-peak with a window of the median beat-to-beat interval, and stacked to form a 2D matrix (number of beats times number of samples of the segmented beats) using the event stacker function from OSET [50], where each row represents one beat. SVD was then applied to this matrix to extract singular values. The resulting values were normalized by the largest singular value and used as features to capture the similarity and reproducibility of ECG beats across the segment. The number of non-zero singular values of a rectangular matrix is smaller than or equal to the minimum of its rows and columns, which in our case was the number of beats used to construct the stacked beat matrix. To ensure a fixed feature length across all subjects and records, the SVD-based feature vector was set to a length of 45, corresponding to the maximum number of beats over 30 s across all subjects. Shorter vectors were zero-padded to reach this length.
viii. *Black-Swan:* This set includes 150 features developed by a top-performing team in the PhysioNet Challenge 2017 for atrial fibrillation classification [49]. The features span multiple domains, including time, frequency, time-frequency, phase space, and meta-level representations. This set has also been successfully applied in other ECG classification tasks [56–58].

The amplitude, interval, and morphological features described above were computed per beat. These beat-wise values were then summarized using the mean, median, and standard deviation to form fixed-length feature vectors.

### 2.5. Machine learning models

Decision tree-based regression models were used for their performance, their ability to handle feature sets with missing values, and their capacity to model complex and nonlinear relationships in data [59]. This includes Extreme Gradient Boosting (XGBoost), Random Forest (RF), CatBoost, and Light Gradient Boosting Machine (LightGBM).

### 2.6. Model developing and data splitting

Our previous studies have shown that, at the population level, males exhibit higher BP than females [39]. Therefore, our SBP and DBP estimation models were trained separately for each sex group. Furthermore, for each sex group, two distinct BP models were trained using: 1) only normal-labeled ECG records and 2) all records. This allowed us to investigate whether BP estimation performance differs when trained exclusively on normal ECGs versus both normal and abnormal cases. As a result, four distinct models were developed for each of SBP and DBP (male-normal/all and female-normal/all). See Table 3 for the breakdown.

All models were trained using 5-fold cross-validation to assess robustness in performance. For training and validation, we used *subject-level* data splitting rather than record-level to avoid inter-subject data leakage between training and validation, ensuring that the results are generalizable to other datasets. The analysis of the pre-processed data showed that the number of ECG-BP pairs per subject varied significantly: over 26% of subjects had only one pair; the median was six pairs per subject; and, in an extreme case, one subject had 3,689 measurement pairs. To address this imbalance and reduce the risk of biasing the ML models toward subjects with more measurements, the number of ECG-BP pairs per subject was capped at six in the train and test datasets. For subjects with more than six recordings, six pairs were randomly selected during cross-validation to make the best use of the available data. Therefore, most ECG-BP pairs eventually contributed to model training and evaluation.

### 2.7. Evaluation metrics

The performance of the developed ML models was evaluated using various metrics, including mean error (ME), standard deviation (SD) of ME, mean absolute error (MAE), SD of MAE, and correlation coefficient, to enable comparison with other studies. Specifically, the correlation coefficient reflects the strength of the linear relationship between the estimated and actual BP values. The correlation coefficient can be either positive or negative, implying a direct or inverse relationship. The absolute correlation coefficient ranges from 0, indicating no linear relationship, to 1, indicating a perfect relationship [60].

## 3. Results

### 3.1. ECG-based BP estimation

Table 4 summarizes the performance of the ML models in estimating DBP and SBP using only ECGs, based on sex and ECG labels (across all and normal-only ECG records).

**Table 4.**
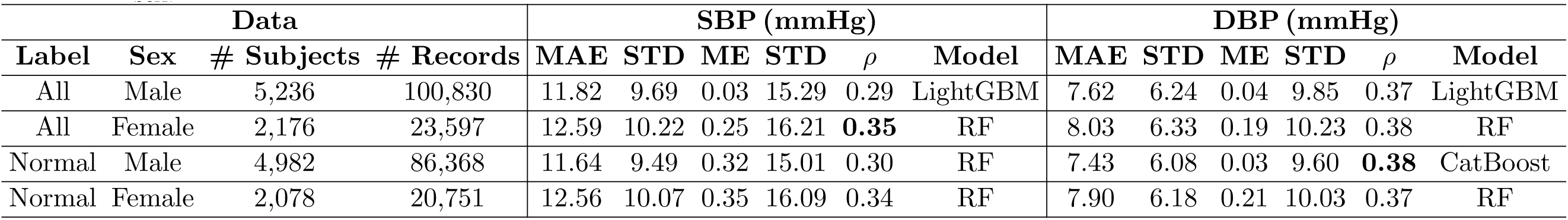
Performance of machine learning models (Random Forest (RF), CatBoost, and Light Gradient Boosting Machine (LightGBM)) for estimating systolic (SBP) and diastolic (DBP) blood pressure using 30 s ECGs with a 280-feature set, split by ECG labels and subject sex.

| Data |  |  |  | SBP (mmHg) |  |  |  |  |  | DBP (mmHg) |  |  |  |  |  |
| --- | --- | --- | --- | --- | --- | --- | --- | --- | --- | --- | --- | --- | --- | --- | --- |
| Label | Sex | # Subjects | # Records | MAE | STD | ME | STD | $\rho$ | Model | MAE | STD | ME | STD | $\rho$ | Model |
| All | Male | 5,236 | 100,830 | 11.82 | 9.69 | 0.03 | 15.29 | 0.29 | LightGBM | 7.62 | 6.24 | 0.04 | 9.85 | 0.37 | LightGBM |
| All | Female | 2,176 | 23,597 | 12.59 | 10.22 | 0.25 | 16.21 | **0.35** | RF | 8.03 | 6.33 | 0.19 | 10.23 | 0.38 | RF |
| Normal | Male | 4,982 | 86,368 | 11.64 | 9.49 | 0.32 | 15.01 | 0.30 | RF | 7.43 | 6.08 | 0.03 | 9.60 | **0.38** | CatBoost |
| Normal | Female | 2,078 | 20,751 | 12.56 | 10.07 | 0.35 | 16.09 | 0.34 | RF | 7.90 | 6.18 | 0.21 | 10.03 | 0.37 | RF |

The best results, based on the correlation coefficient metric, were achieved in estimating DBP with a value of 0.38 using CatBoost and normal-ECG records of males, and in estimating SBP with a value of 0.35 using RF and all-ECG records of females.

Fig. 3 illustrates the results of the prediction errors distribution (the difference between predicted and actual BP values) and 95% percentile contour plots of predicted vs. actual BP for the best-performing estimation models. In an unbiased and well-performing model, the predicted BP values should closely match the actual value, and the prediction errors should have a mean of zero—ideally exhibiting a symmetric unbiased distribution around this mean. However, in Fig. 3, we can see a non-zero mean and skewed error distribution, indicating a systematic bias and asymmetric error.

**Figure 3.**
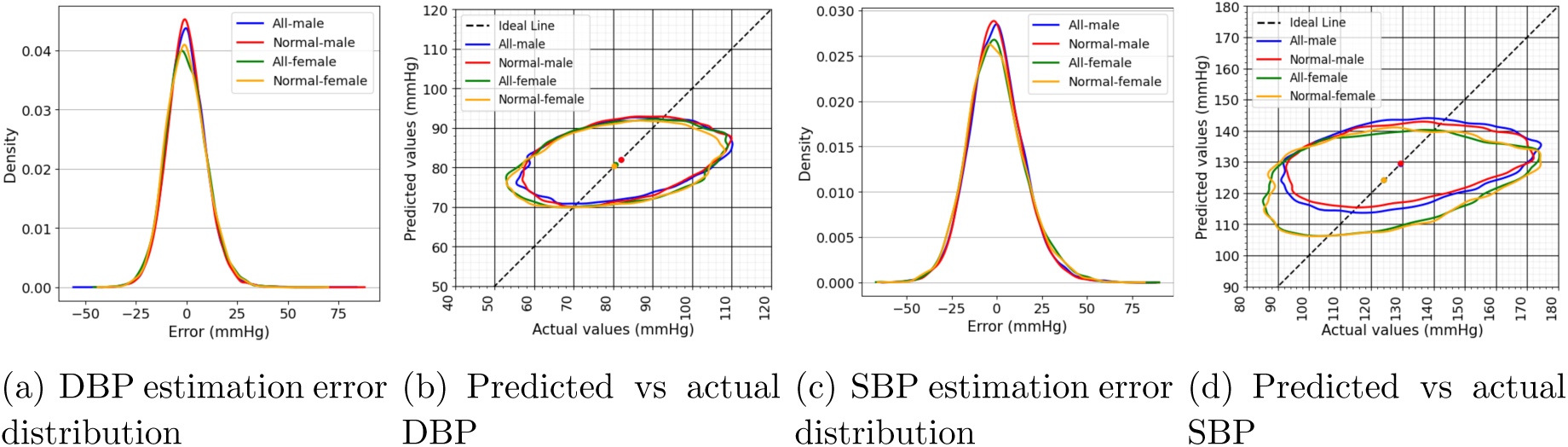
Performance comparison of ML models for estimating systolic (SBP) and diastolic (DBP) blood pressure from 30 s ECGs using 280 features, grouped by ECG label and sex. 3(a) and 3(c) show error PDFs; 3(b) and 3(d), 95% percentile contour plots of predicted vs. actual BP. Dots indicate mean actual and predicted values. For ideal regression, contour plots would be narrow and aligned around the identity line.

The performance of the proposed models was assessed using two widely recognized BP evaluation standards: the Association for the Advancement of Medical Instrumentation (AAMI) and the British Hypertension Society (BHS) [41]. According to the AAMI standard, a valid BP measurement model must achieve a mean error (ME) of *≤*5 mmHg and a standard deviation (SD) of ME *≤*8 mmHg. The BHS standard, in contrast, grades BP measurement devices based on the cumulative percentage of predictions within 5, 10, and 15 mmHg, assigning Grades A, B, or C accordingly. Based on these criteria, neither the SBP nor the DBP prediction models we developed on our dataset satisfied the AAMI requirements, as both exhibited ME and SD values exceeding the thresholds. With respect to the BHS grading, the SBP model failed to meet the standard, while the DBP model fulfills Grade C performance (Table 5).

**Table 5.** Performance of machine learning models (Random Forest (RF), CatBoost, and Light Gradient Boosting Machine (LightGBM)) for estimating systolic (SBP) and diastolic (DBP) blood pressure using 30 s ECGs with a 280-feature set, split by ECG labels and subject sex based on the BHS standard. [**41**]

| BP | Label | Sex | Model | Cumulative Mean Absolute Error<br>Percentage |  |  | # Subjects |
| --- | --- | --- | --- | --- | --- | --- | --- |
| | | | | $\leq 5$ mmHg | $\leq 10$ mmHg | $\leq 15$ mmHg | |
| DBP | All | Male | LightGBM | 44.5% | 73.9% | 89.4% | 5,236 |
| DBP | All | Female | RF | 42.2% | 71.4% | 88.0% | 2,176 |
| DBP | Normal | Male | CatBoost | 46.2% | 74.7% | 89.9% | 4,982 |
| DBP | Normal | Female | RF | 42.4% | 71.5% | 88.4% | 2,078 |
| SBP | All | Male | LightGBM | 30.4% | 53.8% | 71.5% | 5,236 |
| SBP | All | Female | RF | 28.6% | 50.6% | 68.8% | 2,176 |
| SBP | Normal | Male | RF | 30.7% | 54.1% | 72.2% | 4,982 |
| SBP | Normal | Female | RF | 28.5% | 50.6% | 68.7% | 2,078 |
| Grade A | - | - | - | 60% | 85% | 95% | $\geq 85$ |
| Grade B | - | - | - | 50% | 75% | 90% | $\geq 85$ |
| Grade C | - | - | - | 40% | 65% | 85% | $\geq 85$ |

### 3.2. Subject-wise vs. Record-wise Model Training

To evaluate the effect of data partitioning strategies on model performance, we conducted an additional experiment using record-wise cross-validation—where training and validation data were randomly split across individual records, regardless of subject identity. In this setting, the models achieved significantly higher correlation coefficients of 0.59 for SBP and 0.63 for DBP using RF, compared to 0.29 to 0.37 in the subject-wise setup described earlier. This increase in performance suggests that when data splitting is not performed correctly (i.e., using record-wise instead of subject-wise splitting), the models may be leveraging subject-specific patterns seen during training, rather than learning generalizable physiological relationships between ECG and BP that would transfer to unseen subjects.

## 4. Discussion

This study examined the feasibility of estimating BP using only ECG measurements. Despite leveraging a large and diverse dataset, a comprehensive engineered feature set, and robust ML models, the results suggest that ECG-based BP estimation is not practically viable.

### 4.1. Model Performance and Generalization

Model performance on the validation set was low, with correlation coefficients around 0.30, indicating poor generalizability.

Apparently, the models consistently predicted BP values centered around the dataset mean. To explore this, we compared the model outputs with fixed values derived from the training set’s mean, median, and mode. As shown in Table 6, the ML model predictions were nearly indistinguishable from simply using the median BP value, indicating that the models were effectively regressing to the mean.

**Table 6.**
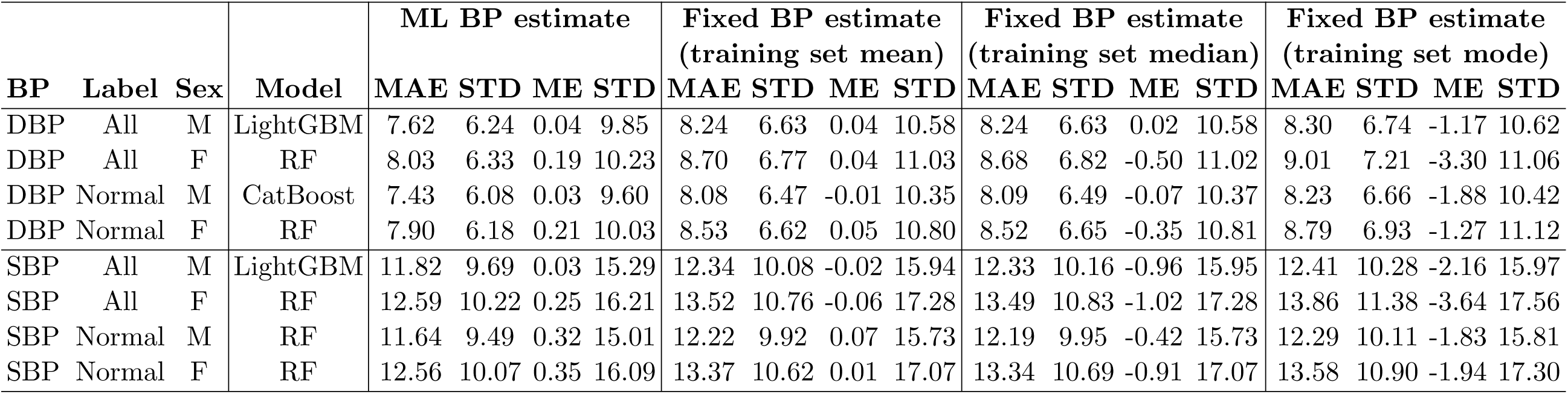
Comparison of best-performing machine learning models (Random Forest (RF), CatBoost, and Light Gradient Boosting Machine (LightGBM)) for estimating systolic (SBP) and diastolic (DBP) blood pressure using 30 s ECGs with a 280-feature set, split by ECG labels and subject sex (Female (F) and male (M)). Fixed values derived from the training set’s mean, median, and mode are included as constant predictions for comparison. (All values are reported in mmHg). Accordingly, ML-based estimates only marginally outperform fixed estimates using population level priors.

This phenomenon, known as *central tendency bias* or *regression to the mean* [61], occurs when a model lacks informative input features. In this case, the model appears to ignore ECG variability and rely instead on the statistical distribution of BP in the training data. Formally, this behavior suggests:

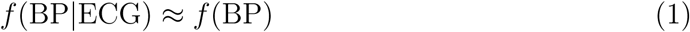

where *f* (*·*) denotes the probability density function, indicating that the ECG features contribute little to the conditional BP distribution.

### 4.2. Comparison with Prior Studies and Standards

Our findings support earlier studies, which debated that accurate ECG-based BP estimation is unfeasible [23, 24], and differ from several earlier studies that reported promising results for ECG-based BP estimation [26–28, 30–38], as summarized in Table 1. Accordingly, many of these prior studies relied on small sample sizes, lacked subject-level separation between training and testing, or used limited feature sets. The current study, using a large ambulatory dataset and rigorous subject-level validation, offers a more generalizable and conservative evaluation.

To assess the potential for clinical utility, we evaluated our models using two widely accepted standards: the AAMI and BHS. None of the models satisfied the AAMI thresholds (mean error *≤* 5 mmHg, SD *≤* 8 mmHg), and only the DBP models marginally achieved Grade C according to the BHS scale. These results further emphasize that ECG-only BP estimation does not meet the performance required for clinical use.

### 4.3. Importance of Across-Subject Validation

The subject-wise versus record-wise partitioning of training and validation records is another aspect often undocumented or overlooked in prior studies. Our results show that record-wise validation can significantly inflate model performance by allowing data from the same subject to appear in both training and test sets. This causes subject-level information leakage, particularly when some individuals contribute many records. As a result, the model learns subject-specific patterns rather than general physiological relationships between ECG and BP, performing well on familiar data but failing to generalize to new subjects. In contrast, our subject-wise approach—where all data from each subject was isolated to either training or validation—prevented this leakage and revealed the true complexity of the task. We also limited the number of records per subject to reduce bias from overrepresented individuals. The lower correlation coefficients in this setup provide a more realistic picture of model performance and reflect the actual difficulty of ECG-based BP estimation.

### 4.4. Physiological and Statistical Implications

From a physiological standpoint, the results can be explained by the fact that BP reflects vascular compliance, peripheral resistance, blood volume, and autonomic tone—factors that are not directly encoded in the electrical activity captured by the ECG. Statistically, this aligns with the concept of *parameter identifiability* in regression problems [62]. Even with a large dataset and highly expressive models, some outputs remain fundamentally non-identifiable from a given input modality. Our findings suggest that BP estimation from ECG alone falls into this category.

### 4.5. Limitations and Future Work

Several limitations should be acknowledged. First, only single-lead ECGs were used, limiting the available morphological and spatial information. Second, ECG and BP were not recorded simultaneously, though the time gap was constrained to a 3.5-minute window and included as a model input. Additionally, all BP values were obtained using non-invasive home devices, which can introduce measurement noise and inaccurate cuff placement and subject positioning.

Future work should investigate models that integrate ECG with additional synchronous physiological signals, such as photoplethysmography (PPG), impedance cardiography, or accelerometry. Multi-lead ECG recordings may also provide more discriminative features. Furthermore, instead of predicting instantaneous BP values, it may be more plausible to estimate average BP over longer time windows. Finally, deep learning models applied directly to ECG waveforms could be explored, although the identifiability limitation observed in this study may still persist.

## 5. Conclusion

This study critically examined the hypothesis that BP can be estimated using only ECG signals. To rigorously test this hypothesis, we developed sex-aware ML regression models using a large ambulatory dataset consisting of 30-second ECG recordings, which are representative of commercially available portable devices. A comprehensive set of 280 engineered ECG features was extracted across time, spatial, and time-spatial domains, and the models incorporated key demographic information such as sex and age. Rigorous data preprocessing and subject-level splitting were applied to minimize bias and ensure generalizability.

Despite these comprehensive modeling strategies, the best-performing models achieved low correlation coefficients between actual and predicted BP values—0.35 for SBP and 0.38 for DBP—indicating limited predictive power. Moreover, the observed prediction performance was comparable to using simple central tendency measures (e.g., the median BP value) as model outputs. These findings suggest that the ECG alone does not carry sufficient information to reliably estimate BP.

In conclusion, while ECG signals remain highly valuable for a wide range of diagnostic applications, their use in isolation for accurate BP estimation is not feasible based on current evidence. Future research should consider combining ECG with other physiological signals or contextual data to improve BP prediction performance or explore alternative applications where ECG-based modeling may yield more robust results.

## Data Availability

No public data

## References

[1] S. S. Mousavi, M. A. Reyna, G. D. Clifford, and R. Sameni, “A Survey on Blood Pressure Measurement Technologies: Addressing Potential Sources of Bias,” Sensors, vol. 24, no. 6, p. 1730, Mar. 2024. [Online]. Available: 10.3390/s24061730

[2] N. WW, “Mcdonald’s blood flow in arteries,” Therorectical, experimental and clinical principles, pp. 54–401, 1998.

[3] T. G. Pickering, J. E. Hall, L. J. Appel, B. E. Falkner, J. Graves, M. N. Hill, D. W. Jones, T. Kurtz, S. G. Sheps, and E. J. Roccella, “Recommendations for Blood Pressure Measurement in Humans and Experimental Animals: Part 1: Blood Pressure Measurement in Humans: A Statement for Professionals From the Subcommittee of Professional and Public Education of the American Heart Association Council on High Blood Pressure Research,” Hypertension, vol. 45, no. 1, p. 142–161, Jan. 2005. [Online]. Available: 10.1161/01.HYP.0000150859.47929.8e

[4] R. Mukkamala, S. G. Shroff, K. G. Kyriakoulis, A. P. Avolio, and G. S. Stergiou, “Cuffless Blood Pressure Measurement: Where Do We Actually Stand?” Hypertension, vol. 82, no. 6, p. 957–970, Jun. 2025. [Online]. Available: 10.1161/HYPERTENSIONAHA.125.24822

[5] P. Muntner, P. T. Einhorn, W. C. Cushman, P. K. Whelton, N. A. Bello, P. E. Drawz, B. B. Green, D. W. Jones, S. P. Juraschek, K. L. Margolis, E. R. Miller, A. M. Navar, Y. Ostchega, M. K. Rakotz, B. Rosner, J. E. Schwartz, D. Shimbo, G. S. Stergiou, R. R. Townsend, J. D. Williamson, J. T. Wright, and L. J. Appel, “Blood Pressure Assessment in Adults in Clinical Practice and Clinic-Based Research,” Journal of the American College of Cardiology, vol. 73, no. 3, p. 317–335, Jan. 2019. [Online]. Available: 10.1016/j.jacc.2018.10.069

[6] P. K. Whelton, R. M. Carey, W. S. Aronow, D. E. Casey, K. J. Collins, C. Dennison Himmelfarb, S. M. DePalma, S. Gidding, K. A. Jamerson, D. W. Jones, E. J. MacLaughlin, P. Muntner, B. Ovbiagele, S. C. Smith, C. C. Spencer, R. S. Stafford, S. J. Taler, R. J. Thomas, K. A. Williams, J. D. Williamson, and J. T. Wright, “2017 ACC/AHA/AAPA/ABC/ACPM/AGS/APhA/ASH/ASPC/NMA/PCNA Guideline for the Prevention, Detection, Evaluation, and Management of High Blood Pressure in Adults,” Journal of the American College of Cardiology, vol. 71, no. 19, p. e127–e248, May 2018. [Online]. Available: 10.1016/j.jacc.2017.11.006

[7] S. Kaplan Berkaya, A. K. Uysal, E. Sora Gunal, S. Ergin, S. Gunal, and M. B. Gulmezoglu, “A survey on ECG analysis,” Biomedical Signal Processing and Control, vol. 43, p. 216–235, May 2018. [Online]. Available: 10.1016/j.bspc.2018.03.003

[8] A. B. De Luna, V. N. Batchvarov, and M. Malik, “The morphology of the electrocardiogram,” The ESC Textbook of Cardiovascular Medicine Blackwell Publishing, vol. 35, 2006.

[9] K. Shah, A. Pandya, P. Kotwani, S. Saha, C. Desai, K. Tyagi, D. Saxena, T. Puwar, and S. Gaidhane, “Cost-Effectiveness of Portable Electrocardiogram for Screening Cardiovascular Diseases at a Primary Health Center in Ahmedabad District, India,” Frontiers in Public Health, vol. 9, Dec. 2021. [Online]. Available: 10.3389/fpubh.2021.753443

[10] L. Neri, M. T. Oberdier, K. C. J. van Abeelen, L. Menghini, E. Tumarkin, H. Tripathi, S. Jaipalli, A. Orro, N. Paolocci, I. Gallelli, M. Dall’Olio, A. Beker, R. T. Carrick, C. Borghi, and H. R. Halperin, “Electrocardiogram Monitoring Wearable Devices and Artificial-Intelligence-Enabled Diagnostic Capabilities: A Review,” Sensors, vol. 23, no. 10, p. 4805, May 2023. [Online]. Available: 10.3390/s23104805

[11] M. A. Muzammil, S. Javid, A. K. Afridi, R. Siddineni, M. Shahabi, M. Haseeb, F. Fariha, S. Kumar, S. Zaveri, and A. J. Nashwan, “Artificial intelligence-enhanced electrocardiography for accurate diagnosis and management of cardiovascular diseases,” Journal of Electrocardiology, vol. 83, p. 30–40, Mar. 2024. [Online]. Available: 10.1016/j.jelectrocard.2024.01.006

[12] S. S. Mousavi, M. Firouzmand, M. Charmi, M. Hemmati, M. Moghadam, and Y. Ghorbani, “Blood pressure estimation from appropriate and inappropriate PPG signals using A whole-based method,” Biomedical Signal Processing and Control, vol. 47, pp. 196–206, 2019.

[13] C. Ma, P. Zhang, F. Song, Y. Sun, G. Fan, T. Zhang, Y. Feng, and G. Zhang, “KD-Informer: A Cuff-Less Continuous Blood Pressure Waveform Estimation Approach Based on Single Photoplethysmography,” IEEE Journal of Biomedical and Health Informatics, vol. 27, no. 5, p. 2219–2230, May 2023. [Online]. Available: 10.1109/JBHI.2022.3181328

[14] S.-S. Lee, D.-H. Nam, Y.-S. Hong, W.-B. Lee, I.-H. Son, K.-H. Kim, and J.-G. Choi, “Measurement of Blood Pressure Using an Arterial Pulsimeter Equipped with a Hall Device,” Sensors, vol. 11, no. 2, p. 1784–1793, Jan. 2011. [Online]. Available: 10.3390/s110201784

[15] D.-H. Nam, W.-B. Lee, Y.-S. Hong, and S.-S. Lee, “Measurement of spatial pulse wave velocity by using a clip-type pulsimeter equipped with a Hall sensor and photoplethysmography,” Sensors, vol. 13, no. 4, pp. 4714–4723, 2013.

[16] Y. Zhang, Y. Li, X. Chen, and N. Deng, “Mechanism of magnetic pulse wave signal for blood pressure measurement,” Journal of Biomedical Science and Engineering, vol. 9, no. 10, pp. 29–36, 2016.

[17] Z. Chen, X. Yang, J. T. Teo, and S. H. Ng, “Noninvasive monitoring of blood pressure using optical Ballistocardiography and Photoplethysmograph approaches,” in 2013 35th annual international conference of the IEEE engineering in medicine and biology society (EMBC). IEEE, 2013, pp. 2425–2428.

[18] C.-S. Kim, A. M. Carek, O. T. Inan, R. Mukkamala, and J.-O. Hahn, “Ballistocardiogram-Based Approach to Cuffless Blood Pressure Monitoring: Proof of Concept and Potential Challenges,” IEEE Transactions on Biomedical Engineering, vol. 65, no. 11, p. 2384–2391, Nov. 2018. [Online]. Available: 10.1109/TBME.2018.2797239

[19] S.-H. Liu, D.-C. Cheng, and C.-H. Su, “A cuffless blood pressure measurement based on the impedance plethysmography technique,” Sensors, vol. 17, no. 5, p. 1176, 2017.

[20] T. H. Huynh, R. Jafari, and W.-Y. Chung, “Noninvasive Cuffless Blood Pressure Estimation Using Pulse Transit Time and Impedance Plethysmography,” IEEE Transactions on Biomedical Engineering, vol. 66, no. 4, p. 967–976, Apr. 2019. [Online]. Available: 10.1109/TBME.2018.2865751

[21] K. Bird, G. Chan, H. Lu, H. Greeff, J. Allen, D. Abbott, C. Menon, N. H. Lovell, N. Howard, W.-S. Chan, R. R. Fletcher, A. Alian, R. Ward, and M. Elgendi, “Assessment of hypertension using clinical electrocardiogram features: A first-ever review,” Frontiers in Medicine, vol. 7, Dec. 2020. [Online]. Available: 10.3389/fmed.2020.583331

[22] S. S. Mousavi, M. Charmi, M. Firouzmand, M. Hemmati, M. Moghadam, and Y. Ghorbani, “ECG-based blood pressure estimation using Mechano-Electric coupling concept,” *arXiv preprint arXiv:2008.10099,* 2020. [Online]. Available: https://arxiv.org/pdf/2008.10099

[23] T. Sato, “Commentary: Assessment of Hypertension Using Clinical Electrocardiogram Features: A First-Ever Review,” Frontiers in Medicine, vol. 8, Jul. 2021. [Online]. Available: 10.3389/fmed.2021.691330

[24] C. Landry and R. Mukkamala, “Current evidence suggests that estimating blood pressure from convenient ECG waveforms alone is not viable,” Journal of Electrocardiology, vol. 81, p. 153–155, Nov. 2023. [Online]. Available: 10.1016/j.jelectrocard.2023.09.001

[25] M. K. B. A. Hassan, M. Y. Mashor, N. F. Mohd Nasir, and S. Mohamed, Measuring of Systolic Blood Pressure Based On Heart Rate. Springer Berlin Heidelberg, 2008, p. 595–598. [Online]. Available: 10.1007/978-3-540-69139-6 149

[26] M. Simjanoska, M. Gjoreski, M. Gams, and A. Madevska Bogdanova, “Non-Invasive Blood Pressure Estimation from ECG Using Machine Learning Techniques,” Sensors, vol. 18, no. 4, p. 1160, Apr. 2018. [Online]. Available: 10.3390/s18041160

[27] M. Simjanoska, S. Kochev, J. Tanevski, A. M. Bogdanova, G. Papa, and T. Eftimov, “Multi-level information fusion for learning a blood pressure predictive model using sensor data,” Information Fusion, vol. 58, p. 24–39, Jun. 2020. [Online]. Available: 10.1016/j.inffus.2019.12.008

[28] F. Miao, B. Wen, Z. Hu, G. Fortino, X.-P. Wang, Z.-D. Liu, M. Tang, and Y. Li, “Continuous blood pressure measurement from one-channel electrocardiogram signal using deep-learning techniques,” Artificial Intelligence in Medicine, vol. 108, p. 101919, Aug. 2020. [Online]. Available: 10.1016/j.artmed.2020.101919

29. A. A. E. Johnson, T. J. Pollard, L. Shen, L.-w. H. Lehman, M. Feng, M. Ghassemi, B. Moody, P. Szolovits, L. Anthony Celi, and R. G. Mark, “MIMIC-III, a freely accessible critical care database,” Scientific Data, vol. 3, no. 1, May 2016. [Online]. Available: 10.1038/sdata.2016.35

[29] S. S. Mousavi, M. Hemmati, M. Charmi, M. Moghadam, M. Firouzmand, and Y. Ghorbani, “Cuff-Less Blood Pressure Estimation Using Only the ECG Signal in Frequency Domain,” in 2018 8th International Conference on Computer and Knowledge Engineering (ICCKE). IEEE, Oct. 2018, p. 147–152. [Online]. Available: 10.1109/ICCKE.2018.8566583

[30] S. Banerjee, B. Kumar, A. P. James, and J. N. Tripathi, “Blood Pressure Estimation from ECG Data Using XGBoost and ANN for Wearable Devices,” in 2022 29th IEEE International Conference on Electronics, Circuits and Systems (ICECS). IEEE, Oct. 2022, p. 1–4. [Online]. Available: 10.1109/ICECS202256217.2022.9970924

[31] C. Wuerich, F. Wichum, O. El-Kadri, K. Ghantawi, N. Grewal, C. Wiede, and K. Seidl, “Blood Pressure Estimation based on Electrocardiograms,” Current Directions in Biomedical Engineering, vol. 8, no. 2, p. 53–56, Aug. 2022. [Online]. Available: 10.1515/cdbme-2022-1015

[32] K. M. Syah, K. N. Pramudia, and M. Rochmad, “Implementation of Artificial Neural Network for Real-Time Blood Pressure Estimation using ECG Signal,” in 2023 10th International Conference on Electrical Engineering, Computer Science and Informatics (EECSI). IEEE, Sep. 2023, p. 12–18. [Online]. Available: 10.1109/EECSI59885.2023.10295886

[33] E. A. Aldein, M. Abdleraheem, U. S. Mohamed, and M. Atef, “An ECG-Based Blood Pressure Estimation Using U-Net auto-encoder and Random Forest Regressor,” in 2023 International Conference on Microelectronics (ICM). IEEE, Dec. 2023, p. 107–112. [Online]. Available: 10.1109/ICM60448.2023.10378899

[34] I. Kuzmanov, E. Zdravevski, P. Lamenski, B. Stojkoska, and A. M. Bogdanova, “A Study on Appropriate Segment Length for Generalized Cuff-less Blood Pressure Estimation from ECG Features,” in 2024 47th MIPRO ICT and Electronics Convention (MIPRO). IEEE, May 2024, p. 1181–1186. [Online]. Available: 10.1109/MIPRO60963.2024.10569523

[35] E. A. Aldein, M. AbdelRaheem, M. M. Abdellatif, U. S. Mohamed, and M. Atef, “ECG-Based Blood Pressure Estimation Using a Two-Stage Inception-Regression Model,” in 2025 International Wireless Communications and Mobile Computing (IWCMC). IEEE, May 2025, p. 914–919. [Online]. Available: 10.1109/IWCMC65282.2025.11059438

[36] X. Fan, H. Wang, Y. Zhao, Y. Li, and K. L. Tsui, “An Adaptive Weight Learning-Based Multitask Deep Network for Continuous Blood Pressure Estimation Using Electrocardiogram Signals,” Sensors, vol. 21, no. 5, p. 1595, Feb. 2021. [Online]. Available: 10.3390/s21051595

[37] X. Fan, H. Wang, F. Xu, Y. Zhao, and K.-L. Tsui, “Homecare-Oriented Intelligent Long-Term Monitoring of Blood Pressure Using Electrocardiogram Signals,” IEEE Transactions on Industrial Informatics, vol. 16, no. 11, p. 7150–7158, Nov. 2020. [Online]. Available: 10.1109/TII.2019.2962546

[38] S. S. Mousavi, Y. Guo, C. Robichaux, A. Sarker, and R. Sameni, “Learning from Two Decades of Blood Pressure Data: Demography-Specific Patterns Across 75 Million Patient Encounters,” in 2024 46th Annual International Conference of the IEEE Engineering in Medicine and Biology Society (EMBC). IEEE, Jul. 2024, p. 1–4. [Online]. Available: 10.1109/EMBC53108.2024.10781724

[39] M. L. Munoz, A. van Roon, H. Riese, C. Thio, E. Oostenbroek, I. Westrik, E. J. C. de Geus, R. Gansevoort, J. Lefrandt, I. M. Nolte, and H. Snieder, “Validity of (Ultra-)Short Recordings for Heart Rate Variability Measurements,” PLOS ONE, vol. 10, no. 9, p. e0138921, Sep. 2015. [Online]. Available: 10.1371/journal.pone.0138921

[40] G. S. Stergiou, B. Alpert, S. Mieke, R. Asmar, N. Atkins, S. Eckert, G. Frick, B. Friedman, T. Graßl, T. Ichikawa, J. P. Ioannidis, P. Lacy, R. McManus, A. Murray, M. Myers, P. Palatini, G. Parati, D. Quinn, J. Sarkis, A. Shennan, T. Usuda, J. Wang, C. O. Wu, and E. O’Brien, “A universal standard for the validation of blood pressure measuring devices: Association for the Advancement of Medical Instrumentation/European Society of Hypertension/International Organization for Standardization (AAMI/ESH/ISO) Collaboration Statement,” Journal of Hypertension, vol. 36, no. 3, p. 472–478, Mar. 2018. [Online]. Available: 10.1097/HJH.0000000000001634

[41] G. Mancia, A. Ferrari, L. Gregorini, G. Parati, G. Pomidossi, G. Bertinieri, G. Grassi, M. di Rienzo, A. Pedotti, and A. Zanchetti, “Blood pressure and heart rate variabilities in normotensive and hypertensive human beings.” Circulation Research, vol. 53, no. 1, p. 96–104, Jul. 1983. [Online]. Available: 10.1161/01.res.53.1.96

[42] S. L. Graham, S. M. Drance, K. Wijsman, G. R. Douglas, and F. S. Mikelberg, “Ambulatory Blood Pressure Monitoring in Glaucoma,” Ophthalmology, vol. 102, no. 1, p. 61–69, Jan. 1995. [Online]. Available: 10.1016/S0161-6420(95)31053-6

[43] K. Kario, T. G. Pickering, Y. Umeda, S. Hoshide, Y. Hoshide, M. Morinari, M. Murata, T. Kuroda, J. E. Schwartz, and K. Shimada, “Morning Surge in Blood Pressure as a Predictor of Silent and Clinical Cerebrovascular Disease in Elderly Hypertensives: A Prospective Study,” Circulation, vol. 107, no. 10, p. 1401–1406, Mar. 2003. [Online]. Available: 10.1161/01.CIR.0000056521.67546.AA

[44] D. L. Clement, M. L. De Buyzere, D. A. De Bacquer, P. W. de Leeuw, D. A. Duprez, R. H. Fagard, P. J. Gheeraert, L. H. Missault, J. J. Braun, R. O. Six, P. Van Der Niepen, and E. O’Brien, “Prognostic Value of Ambulatory Blood-Pressure Recordings in Patients with Treated Hypertension,” New England Journal of Medicine, vol. 348, no. 24, p. 2407–2415, Jun. 2003. [Online]. Available: 10.1056/NEJMoa022273

[45] L. E. Okamoto, A. Gamboa, C. Shibao, B. K. Black, A. Diedrich, S. R. Raj, D. Robertson, and I. Biaggioni, “Nocturnal Blood Pressure Dipping in the Hypertension of Autonomic Failure,” Hypertension, vol. 53, no. 2, p. 363–369, Feb. 2009. [Online]. Available: 10.1161/HYPERTENSIONAHA.108.124552

[46] F. Sayk, C. Teckentrup, C. Becker, D. Heutling, P. Wellhöner, H. Lehnert, and C. Dodt, “Effects of selective slow-wave sleep deprivation on nocturnal blood pressure dipping and daytime blood pressure regulation,” American Journal of Physiology-Regulatory, Integrative and Comparative Physiology, vol. 298, no. 1, p. R191–R197, Jan. 2010. [Online]. Available: 10.1152/ajpregu.00368.2009

[47] G. Mancia, “Short- and Long-Term Blood Pressure Variability: Present and Future,” Hypertension, vol. 60, no. 2, p. 512–517, Aug. 2012. [Online]. Available: 10.1161/HYPERTENSIONAHA.112.194340

[48] M. Zabihi, A. Bahrami Rad, A. K. Katsaggelos, S. Kiranyaz, S. Narkilahti, and M. Gabbouj, “Detection of Atrial Fibrillation in ECG Hand-held Devices Using a Random Forest Classifier,” in 2017 Computing in Cardiology Conference (CinC), ser. CinC2017. Computing in Cardiology, Sep. 2017. [Online]. Available: 10.22489/CinC.2017.069-336

[49] R. Sameni, The Open-Source Electrophysiological Toolbox (OSET), version 4.0, 2006–2025. [Online]. Available: https://github.com/alphanumericslab/OSET

[50] J. Leski, “Robust weighted averaging [of biomedical signals],” IEEE Transactions on Biomedical Engineering, vol. 49, no. 8, p. 796–804, Aug. 2002. [Online]. Available: 10.1109/TBME.2002.800757

[51] G. Clifford, F. Azuaje, and P. McSharry, Advanced methods and tools for ECG data analysis, ser. Artech House engineering in medicine & biology series. Artech House, 2006.

[52] S. Luo, K. Michler, P. Johnston, and P. W. Macfarlane, “A comparison of commonly used QT correction formulae: The effect of heart rate on the QTc of normal ECGs,” Journal of Electrocardiology, vol. 37, p. 81–90, Oct. 2004. [Online]. Available: 10.1016/j.jelectrocard.2004.08.030

[53] Y. N. Fuadah and K. M. Lim, “Optimal Classification of Atrial Fibrillation and Congestive Heart Failure Using Machine Learning,” Frontiers in Physiology, vol. 12, Feb. 2022. [Online]. Available: 10.3389/fphys.2021.761013

[54] L. Zheng, Z. Wang, J. Liang, S. Luo, and S. Tian, “Effective compression and classification of ECG arrhythmia by singular value decomposition,” Biomedical Engineering Advances, vol. 2, p. 100013, Dec. 2021. [Online]. Available: 10.1016/j.bea.2021.100013

[55] A. Bahrami Rad, C. Galloway, D. Treiman, J. Xue, Q. Li, R. Sameni, D. Albert, and G. D. Clifford, “Atrial fibrillation detection in outpatient electrocardiogram monitoring: An algorithmic crowdsourcing approach,” PLOS ONE, vol. 16, no. 11, p. e0259916, Nov. 2021. [Online]. Available: 10.1371/journal.pone.0259916

[56] A. Bahrami Rad, M. Kirsch, Q. Li, J. Xue, R. Sameni, D. Albert, and G. D. Clifford, “A Crowdsourced AI Framework for Atrial Fibrillation Detection in Apple Watch and Kardia Mobile ECGs,” Sensors, vol. 24, no. 17, p. 5708, Sep. 2024. [Online]. Available: 10.3390/s24175708

[57] Z. Koscova, A. B. Rad, S. Nasiri, M. A. Reyna, R. Sameni, L. M. Trotti, H. Sun, N. Turley, K. L. Stone, R. J. Thomas, E. Mignot, B. Westover, and G. D. Clifford, “From Sleep Patterns to Heart Rhythms: Predicting Atrial Fibrillation from Overnight Polysomnograms,” Journal of Electrocardiology, Jun. 2024. [Online]. Available: 10.1101/2024.06.04.24308444

[58] V. Podgorelec, P. Kokol, B. Stiglic, and I. Rozman, “Decision trees: an overview and their use in medicine,” Journal of medical systems, vol. 26, pp. 445–463, 2002.

[59] J. Martin Bland and D. Altman, “Statistical methods for assessing agreement between two methods of clinical measurement,” The Lancet, vol. 327, no. 8476, p. 307–310, Feb. 1986. [Online]. Available: 10.1016/S0140-6736(86)90837-8

[60] A. G. Barnett, “Regression to the mean: what it is and how to deal with it,” International Journal of Epidemiology, vol. 34, no. 1, p. 215–220, Aug. 2004. [Online]. Available: 10.1093/ije/dyh299

62. [61] R. Sameni, “Beyond Convergence: Identifiability of Machine Learning and Deep Learning Models,” 2023. [Online]. Available: https://arxiv.org/abs/2307.11332

